# LocusBlend: Flexible multi-index regional visualization of genomic association signals

**DOI:** 10.64898/2026.07.15.26358129

**Authors:** Chenyu Yang, Noah Cook, Youjie Zeng, Tianyi Fu, John Budde, Carlos Cruchaga, Michael E. Belloy

## Abstract

**Summary:** It has become standard practice to visualize regional signals from genome-wide association studies (GWAS) using LocusZoom plots. Similarly, GWAS signals are compared to regionally matched quantitative trait loci (QTLs), i.e. variant-to-gene regulation data, using LocusCompare plots to aid assessment of candidate trait-related genes. Despite broad usage, these tools annotate variants by linkage disequilibrium (LD) to a single lead or index variant. This single-index representation has limitations for visualizing complex loci that contain multiple independent signals. We present LocusBlend, an interactive web application for multi-index LD-blended visualization of genomic loci. LocusBlend supports one or two genomic association summary-statistic datasets and one to three index variants, multi-index LocusZoom color-blended plots, and matching LocusCompare visualizations. Applications to Alzheimer’s disease GWAS and QTL signals illustrate LocusBlend enables visualization and separation of independent signals despite shared LD and high genomic complexity. Overall, LocusBlend is aimed at supporting researchers handle the continuously expanding complexity of human genomics findings.

**Availability and Implementation:** LocusBlend is freely available at https://locusblend.wustl.edu. Publication ready plots are generated in <1min. Source code, documentation, example datasets, input templates, and reproducibility instructions are available at https://github.com/Belloy-Lab/LocusBlend. LocusBlend is implemented in Python using Streamlit, Plotly, and PLINK.

**Supplementary Information:** Supplementary data are available at Bioinformatics online.

## Introduction

Genome-wide association studies (GWAS) have identified tens of thousands of loci associated with complex traits and diseases. These loci require close inspection to verify robustness of the association signals and review of the local genomic structure. Moreover, interpretation of these loci remains challenging because most associated variants are non-coding, correlated through linkage disequilibrium (LD), and often located nearby multiple plausible target genes(Visscher *et al*. 2017, Li and Ritchie 2021). Therefore, regional or locus-level visualization has become an important step in GWAS follow-up, allowing researchers to inspect the strength of association signals, local LD structure, nearby genes, recombination patterns, and even overlap with regionally matched molecular quantitative trait loci (QTLs) to help elucidate the molecular mechanisms and genes that may be affected by the GWAS signals. Tools such as LocusZoom, LocusZoom.js, locuszoomr, LocuscCompare, PheWeb, and related regional browsers have made locus-level visualization broadly accessible for static, interactive, and web-based exploration of genetic association results(Pruim *et al*. 2010, Liu *et al*. 2019, Gagliano Taliun *et al*. 2020, Boughton *et al*. 2021, Lewis and Wang 2024).

A common feature of these regional visualization tools is that variants are annotated by linkage disequilibrium (LD) to a single lead or index variants. This is useful for simple loci containing a single independent association signal, but as human genetic studies have become increasingly larger and more powerful, so has the complexity of the genetic signals they pinpoint. This complexity is also increasingly common in post-GWAS interpretation, where GWAS signals are compared with transcript expression (e)QTL, transcript splicing (s)QTL, protein (p)QTL, and other molecular QTL datasets to prioritize candidate genes or tissues (Giambartolomei *et al*. 2014, 2018, Hormozdiari *et al*. 2016, Aguet *et al*. 2017, Wallace 2021, Pullin and Wallace 2025). In this area, LocusCompare plots and other genetic colocalization visualization tools help compare association patterns across two datasets, but they similarly rely on single-index LD visualization, making it difficult to distinguish multiple nearby independent signals (Liu *et al*. 2019). Moreover, many efforts now integrate GWAS with QTL data to perform e.g. transcriptome-wide association studies (TWAS) or protein-wide association studies (PWAS). These can provide new insights into potential trait-causal genes and importantly increase power for gene discovery when multiple independent gene regulatory signals exist at a given locus. As a result, however, these approaches can tease out complex genomic signals that reside below common genome-wide significance (P<5e-8), further emphasizing the need for new visualization strategies.

To fill in this gap, we built LocusBlend, a flexible, interactive web-based approach for multi-index LD-blended visualization of complex genomic loci. LocusBlend extends standard LocusZoom and LocusCompare plots by allowing users to specify up to three index variants within a locus. To showcase its utility, we apply LocusBlend to novel locus discoveries in Alzheimer’s disease (AD) enabled through female-stratified PWAS (Cook *et al*. 2025). We specifically focus on the *PSEN1* and *CD84* regions, where multiple AD GWAS, AD PWAS, and QTL signals overlapped, and LocusBlend enablied visualization and separation of independent signals despite shared LD and high genomic complexity.

### Implementation

#### Overview

LocusBlend is a Python-based web application for interactive locus-level visualization of GWAS and molecular QTL summary statistics, allowing flexible and customizable generation of locus zoom and locus compare plots in a publication ready format. Instead of assigning each variant LD information and color formatting based on only one lead variant, LocusBlend encodes multi-index LD tagging patterns using blended color profiles, while marker size represents LD strength (**Figure 1**).

**Figure 1.**
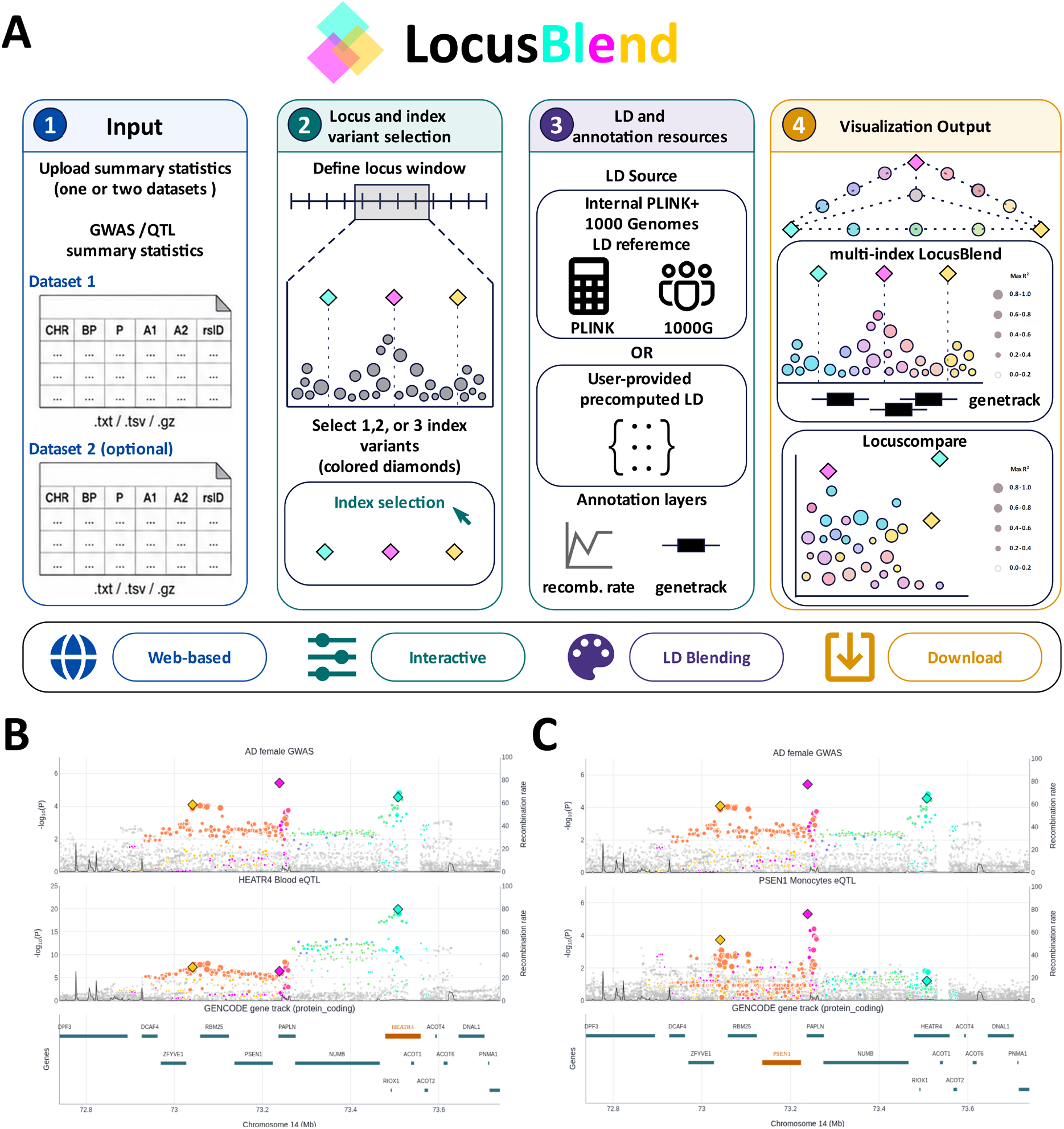
LocusBlend workflow and example output. **A)** LocusBlend uses one or two GWAS or QTL summary-statistic datasets allowing users to select a locus and one to three index variants, using either default PLINK-derived 1000 Genomes LD or user-provided LD. The visualization output includes standard locus zoom or multi-index LD-blended locus zoom and locus compare plots. In the LD-blended plots, diamonds mark index variants, point colors mark multi-index LD patterns with regard to the index variants, and point size reflects maximal LD strength with regard to the index variants. **B)** Example LocusBlend plot at the female-biased AD genetic risk locus *PSEN1*, marking overlap of an AD genetic signal with a blood eQTL for *HEART4* and **C)** for another independent AD genetic signal with a monocyte eQTL for *PSEN1*. Extended plots are provided in SFigure 1A. **Alt text:** Overview of the LocusBlend workflow, showing input datasets, locus and index-variant selection, LD and annotation resources, and example output plots.

The workflow includes uploading one or two genomic summary-statistic datasets, selecting a locus and one to three index variants, selecting LD source data, and finally generating multi-index LD-blended visualizations.

#### Input data and analysis modes

LocusBlend supports two primary analysis modes depending on the number of summary-statistic datasets uploaded. In the one-dataset mode, users provide a single GWAS or QTL summary-statistic file and generate an interactive locus zoom plot for a selected locus. In the two-dataset mode, users provide two summary-statistic files and generate both interactive locus zoom and locus compare plots. When two datasets are provided, LocusBlend allows generating locus compare plots either as separate panels for each selected index variant or as a single panel that encodes LD relationships to multiple index variants simultaneously.

#### Index variant selection and LD calculation

LocusBlend allows users to specify one to three index variants within a selected genomic region. When single index mode is selected, LocusBlend generates a standard locus zoom plot in which variants are annotated according to their LD with the selected index variant. When two or three index variants are provided, LocusBlend switches to a multi-index mode in which LD relationships to all selected index variants are integrated simultaneously. LD information can be obtained in two ways. First, by default, LocusBlend calculates LD internally using PLINK and an embedded 1000 Genomes reference panel (Purcell *et al*. 2007, Auton *et al*. 2015, Chang *et al*. 2015). Second, users can upload precomputed LD values when available.

A core feature of LocusBlend is its multi-index LD-blended visualization. Each index variant is assigned a distinct color profile using standard six-character hexadecimal color codes. Blended colors are generated according to RGB color-mixing principles, such that variants primarily linked to one index variant retain the corresponding color, while variants showing LD to both index variants receive intermediate blended colors. In the three-index mode, the same principle is extended to three index variants, allowing variants to represent signal-specific, shared, or partially overlapping LD relationships within the same plot.

### Application to complex AD loci

We applied LocusBlend to two complex AD loci to demonstrate how multi-index LD-blended visualization can support interpretation and improve visualization of local GWAS and QTL signals. This prior study specifically conducted sex-stratified GWAS and PWAS of AD using brain pQTL data, supplemented by multi-tissue multi-QTL genetic colocalization analyses, to identify sex-biased genetic risk signals and candidate causal genes for AD (Cook *et al*. 2025). We focus on two regions, *PSEN1* and *CD84*, that were identified from the female-stratified AD PWAS and were marked by complex association patterns, including multiple independent AD association signals below conventional genome-wide significance, and overlapping (i.e. colocalizing) molecular QTL signals.

#### PSEN1

The female AD GWAS signals that underlied the PSEN1 PWAS finding spanned three nearby signals that respectively overlapped with several QTL signals, including brain PSEN1 and PPAIP1 pQTLs, a monocyte *PSEN1* eQTL, a dorsolateral prefrontal cortex methylation QTL, and a blood eQTL for *HEATR4* (**Figure 1B-C, SFigure 1A**).

While these signals were challenging to resolve at first, guided by the QTL lead variants and visualization through LocusBlend, it became apparent that the locus contained 3 likely independent AD genetic signals that share LD with one another.

#### CD84

The female AD GWAS signals that underlied the CD84 PWAS findings spanned 4 independent signals that respectively overlapped with several QTL signals, including CSF CD84 and FCER1G pQTLs, microglial *FCER1G* eQTLs, monocyte *FCER1G* eQTLs, and a blood eQTL for *NDUFS2* (**SFigure 1B**). Using LocusBlend, it became apparent that 2 of these 4 signals drove the CD84 PWAS finding, while the other 2 pertained to additional AD GWAS signals that overlapped with other gene QTLs.

### Availability, reproducibility, and limitations

LocusBlend is freely available for non-commercial use at https://locusblend.wustl.edu. The source code, together with documentation, example datasets, input templates, and reproducibility instructions, is available in the LocusBlend GitHub repository at https://github.com/Belloy-Lab/LocusBlend. The web application does not require mandatory user registration, and the source code can be deployed locally.

It is important for users to remain aware that LocusBlend is a visualization framework, not a statistical fine-mapping, conditional analysis, or colocalization method. The primary goal of LocusBlend is to help users visualize complex loci and, in turn, guide exploratory identification of candidate independent signals that remain to be verified by additional, complimentary analyses.

## Data availability

Example datasets, input templates, and files required to reproduce the LocusBlend demonstration analyses are available in the LocusBlend GitHub repository at https://github.com/Belloy-Lab/LocusBlend. These example files are provided to demonstrate the expected input format and reproduce the visualizations shown in this manuscript. The complete Alzheimer’s disease sex-stratified GWAS summary statistics used as source data for the AD examples are described in (Cook *et al*. 2025) and should be accessed according to the data availability statement of that study. QTL summary statistics used in the example datasets are available from the original studies. (cf. (Cook *et al*. 2025))

## Code availability

The code used to support the findings of this study is publicly available at GitHub: https://github.com/Belloy-Lab/LocusBlend.

## Author contributions

C.Y.: Conceptualization, Methodology, Software, Formal analysis, Visualization, Writing—original draft. N.C.: Data curation, Formal analysis, Writing—review and editing. Y.Z.: Data curation, Writing—review and editing. T.F.: Data curation, Writing—review and editing. M.E.B.: Conceptualization, Methodology, Supervision, Funding acquisition, Writing—original draft, Writing—review and editing.

## Acknowledgements

We thank the open-source scientific software community for developing and maintaining tools that supported LocusBlend. We are grateful to Prof. Carlos Cruchaga and the Washington University NeuroGenomics and Informatics Center for institutional support, to John Budde for technical support with web deployment and server configuration, and to Washington University Marketing & Communications and Washington University Information Technology for assistance with review, approval, and implementation of the LocusBlend domain.

We thank members at the Washington University NeuroGenomics and Informatics Center for feedback on the LocusBlend interface, documentation, example datasets, and visualization features.

