## Supplement for "LocusBlend: Flexible multi-index regional visualization of genomic association signals"

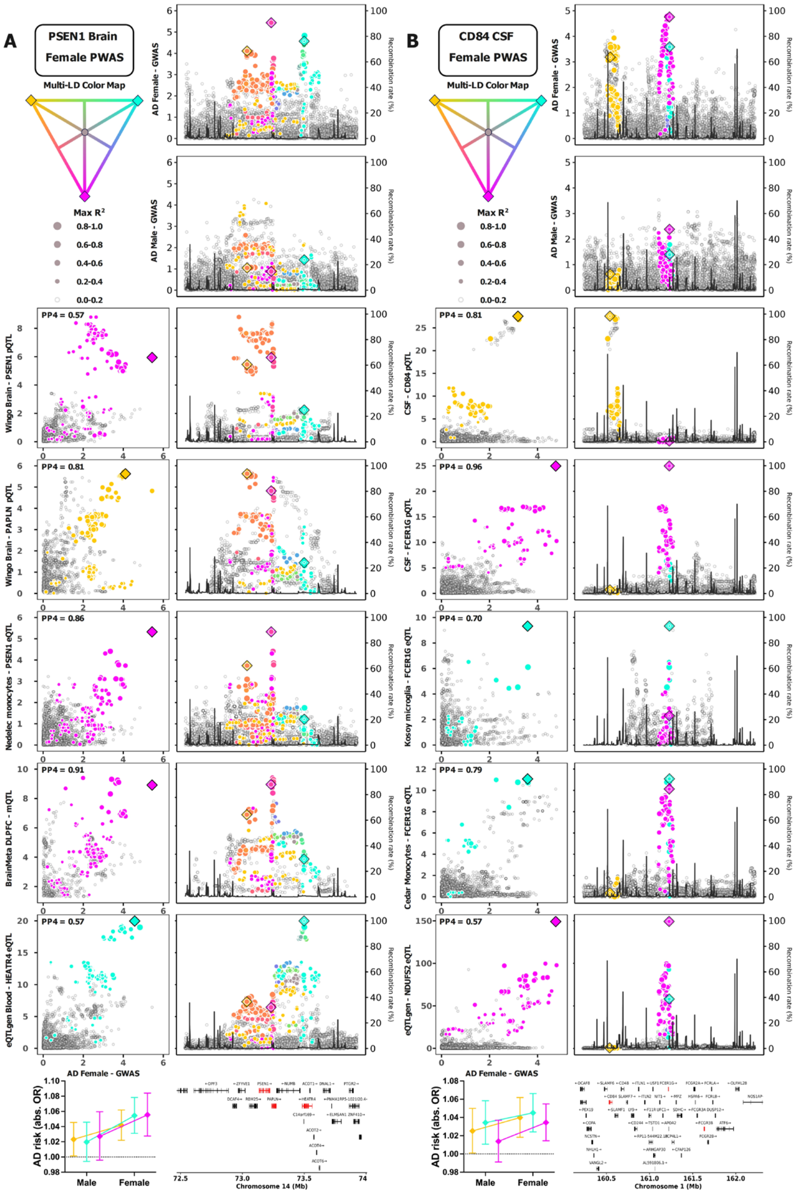


**Supplementary Figure 1. Application of LocusBlend to complex Alzheimer's disease loci identified from female-stratified proteome-wide association studies.** LocusBlend visualizations are shown for **A)** the PSEN1 region and **B)** the CD84 region. For each locus, the multi-LD color map shows three selected index variants and their blended color profiles. Colored points indicate variants in LD with one or more index variants; color represents the multi-index LD tagging pattern, point size represents maximum LD strength, and diamond markers indicate index variants. The panels show sex-stratified AD GWAS signals stacked on top of representative GWAS-QTL comparisons, regional QTL association patterns, and gene annotations. Left top corners show genetic colocalization support (PP4 values ranging 0-1). Left bottom panels show AD risk estimates for index variants across males and females. Altogether, these figures illustrate how LocusBlend enables resolving signals that are (i) specific, (ii) shared, or (iii) have partially overlapping LD patterns across complex GWAS and QTL loci. These data match analyses presented in Cook et al. 2025, for additional, please refer to https://doi.org/10.1101/2025.10.31.25339089.
